# Digitising HIV Testing Registers to Support National Scale-Up of Three-Test HIV Diagnostic Algorithm: A Case Study of an AI-Powered Monitoring and Evaluation System in Malawi

**DOI:** 10.64898/2026.01.08.26343708

**Authors:** Tiwonge Chimpandule, Hannock Tweya, Stone Mbiriyawanda, Tobias Masina, Khumbo Namachapa, Martha Muyaso, Elsie Kasambwe, Chimwemwe Mkandawire, William Wu, Jiehua Chen, Michał Łazowik, Maciej Pomykała, Watipa Nyangulu, Chatonda Ngwira, Maria Sanena, Joyce Mtambo, Molly Ndisale, Dominik Bilicki, Leah Goeke, Confidence Banda, Jeffrey W. Imai-Eaton, Cheryl Case Johnson, Andreas Jahn

**Affiliations:** Directorate of HIV, STI and Viral Hepatitis, Ministry of Health, Lilongwe, Malawi; I-TECH Malawi, Lilongwe, Malawi; Quantitative Engineering Design (QED.ai), Lilongwe, Malawi; Quantitative Engineering Design (QED.ai), Warsaw, Poland; Quantitative Engineering Design (QED.ai), Sheridan, United States; University of North Carolina Project, Malawi-Tidziwe Research Centre, Lilongwe, Malawi; Center for Communicable Disease Dynamics, Department of Epidemiology, Harvard T.H. Chan School of Public Health, Boston, United States; MRC Centre for Global Infectious Disease Analysis, School of Public Health, Imperial College London, London, United Kingdom; Global HIV, Hepatitis and STI Programmes, World Health Organization, Geneva, Switzerland; Department of Global Health and I-TECH, University of Washington, Seattle, United States

**Keywords:** digital system, HIV testing, three-test HIV algorithm, Malawi

## Abstract

**Background:** By November 2022, Malawi became one of the first countries to implement the 2019 World Health Organization (WHO) recommended three-test HIV algorithm nationally. To support the scale-up, a new monitoring and evaluation (M&E) system, ScanForm, was introduced to digitise individual-level records from paper-based HIV testing registers using artificial intelligence (AI). We describe the national scale-up of the ScanForm M&E system alongside the three-test algorithm and assess the performance of the M&E system in monitoring quality assurance and generating routine program reporting.

**Methods:** We conducted a descriptive study using routinely collected HIV testing data captured through ScanForm between November 2022 and September 2025. HIV testing providers photographed completed handwritten register pages, which were automatically transcribed using AI, validated and summarised into reports. The system performance was assessed by evaluating changes over time in numbers of recording errors and HIV testing services (HTS) protocol deviations detected in the first image submitted per register page, data completeness, and reporting timeliness. Trends in recording errors and protocol deviations were analysed among facilities with at least 24 months of follow-up, using each facility’s activation date as the baseline.

**Results:** By January 2023, 260 (30%) of 867 HTS facilities had adopted the M&E system and the three-test algorithm. By September 2024, 98% coverage nationwide was achieved (853/867 facilities). During the study period, 9,082,891 HTS encounters were captured through ScanForm and HIV positivity was 1.8%. A total of 1,412,133 errors and deviations were identified, representing an overall error rate of 0.6%. Of these, 1,370,002 (97%) were recording errors and 42,131 (3%) were deviations in testing protocol. Only two types of deviations were specific to the implementation of the three-test HIV algorithm: 15,051 (36%) were deviations from the HTS testing algorithm, and 4,683 (11%) were misclassifications of HIV test results. Errors and deviations declined by 32% in the first three months and by 58% over 24 months, while the error rate dropped from 1.6% to 0.4% over 24 months. After resolving the validation checks, 99.8% of all mandatory data elements were complete. Approximately 94.6% of all HTS records were submitted on time.

**Conclusion:** The national rollout of the AI-powered M&E system was rapid, achieving national coverage within two years, and the system provided timely and complete program data. Errors and deviations declined during implementation, indicating improved data quality and adherence to the HTS protocol. Integrating digital data systems into routine service delivery has the potential to strengthen guideline implementation and enhance the quality of HIV services at scale.

## Background

HIV Testing Services (HTS) are a crucial entry point for treatment, initiation, and prevention interventions, and play a key role in achieving the global goal of ending AIDS by 2030 [1,2]. Although HIV tests are generally reliable, studies show that challenges related to misdiagnosis or misclassification errors persist [3,4]. Reported misdiagnosis rates average 0.4% for false-negative and 3.1% for false-positive results ([4] Misdiagnosis can arise from assay performance failure or programmatic issues such as recording errors, limited quality assurance, insufficient staff training, inadequate supervision, and suboptimal test storage or transport conditions ([5]. The consequences of misdiagnosis are substantial. At the individual level, misdiagnosis can lead to stigma, psychological trauma, and inappropriate or delayed treatment [6,7], while at the program level, it can lead to resource misallocation. To reduce the risk of misdiagnosis, the World Health Organisation (WHO) recommends a three-test HIV algorithm [8].

By 2022, Malawi became one of the first countries to adopt the 2019 WHO-recommended three-test HIV algorithm [9]. Core HTS quality measures were maintained, including post-marketing kit evaluations, routine facility quality control, biannual provider proficiency testing, and regular site and provider certification [9]. However, traditional in-person supervision, which required national and district personnel to travel to facilities, was costly and not feasible at scale. National scale-up of the three-test algorithm required an M&E system capable of providing near-real-time, individual-level data to support monitoring of adherence to HIV testing protocols, rapid decision-making and targeted interventions.

The existing paper-based monitoring and evaluation (M&E) system could not meet program needs. It produced only aggregated data, which facilities generated by manually tabulating age- and gender- disaggregated monthly reports. Compiled paper reports were collected and entered at district level into District Health Information System (DHIS2) and at national level into the Department of HIV/AIDS Management Information System (DHAMIS). This process was laborious and prone to errors and delays. DHIS2 is the Malawi Ministry of Health’s (MoH) national platform for routine aggregated health service data. Between 2016 and 2022, monthly HTS reporting often met or exceeded the 85% completeness target at national level, but timeliness lagged in most years, reflecting persistent delays in data availability for program monitoring and decision-making. [10]. At the same time, deploying a nationwide electronic medical records system (EMRS) was not feasible due to unreliable infrastructure, including inadequate equipment, limited internet connectivity, and frequent power outages [11–13]. Similar to many other countries in sub-Saharan Africa (SSA), access to electricity remains limited in Malawi; only 15.6% of the population has electricity [14] and scheduled power outages can last up to 10 hours per day [15]. These prolonged outages would force health facilities to maintain both paper-based and point-of-care electronic systems, further limiting EMRS usability.

To address these limitations, Malawi adopted an artificial intelligence (AI)-powered M&E system, ScanForm, developed by Quantitative Engineering Design (QED) [16], to complement in-person supervision visits and provide quality assurance oversight across health facilities. HTS providers completed scannable register pages, which were captured using smartphones and uploaded to a central server where individual-level data were automatically transcribed. ScanForm applied automated validation checks to identify potential recording errors and deviations from the HTS protocol. Validation check results and monthly program reports were automatically shared via smartphones at clinics and on a national web portal for resolution, while monthly program reports were transmitted to the national DHIS2 instance and DHAMIS. By providing feedback on data quality and adherence to the three-test algorithm, the system was expected to help HTS providers improve their recording and implementation practices. Understanding ScanForm’s performance in monitoring quality assurance, acceptability among HTS providers, and implementation costs is essential to inform future adoption and sustainability.

This study assesses the performance of the ScanForm M&E system for monitoring quality assurance and generating routine program reporting. Specifically, we aimed to: (1) Determine the coverage of the M&E system and three-test HIV algorithm; 2) Assess volumes of recording errors and HTS protocol deviations detected in the first image submitted per register page and whether they decreased over time; 3) Assess data completeness and reporting timeliness; 4) Estimate the cost of the M&E system; 5) Examine usability and acceptability of the M&E system.

## Methods

### Study design

This descriptive study analysed routinely collected HTS data from 867 health facilities in Malawi using the ScanForm M&E system between November 2022 and September 2025, complemented by qualitative data from the pilot phase, between March 2021 and December 2021.

### HIV testing services in Malawi

HTS in Malawi are delivered in health facilities and community settings by trained lay and clinical providers. During HIV testing (an HTS encounter), clients receive pre-test counseling, HIV testing, and post-test counseling. The Malawi HTS program uses Determine (Test 1 = T1), Uni-Gold (Test 2 = T2) and Bioline (Test 3 = T3) tests. A non-reactive T1 indicates an HIV-negative result (**Fig 1**). If T1 is reactive, T2 is performed. A reactive T2 leads to T3 where a reactive T3 confirms HIV-positive status; a non-reactive T3 is inconclusive. If T2 is non-reactive, T1 is repeated; a non-reactive result confirms HIV negative status, a reactive result is inconclusive. Inconclusive results require repeating the algorithm after two weeks; persistent inconclusive results prompt PCR testing. When T1 is reactive during community outreach testing, the client is referred to a static facility for all three HIV tests.

**Fig 1.**
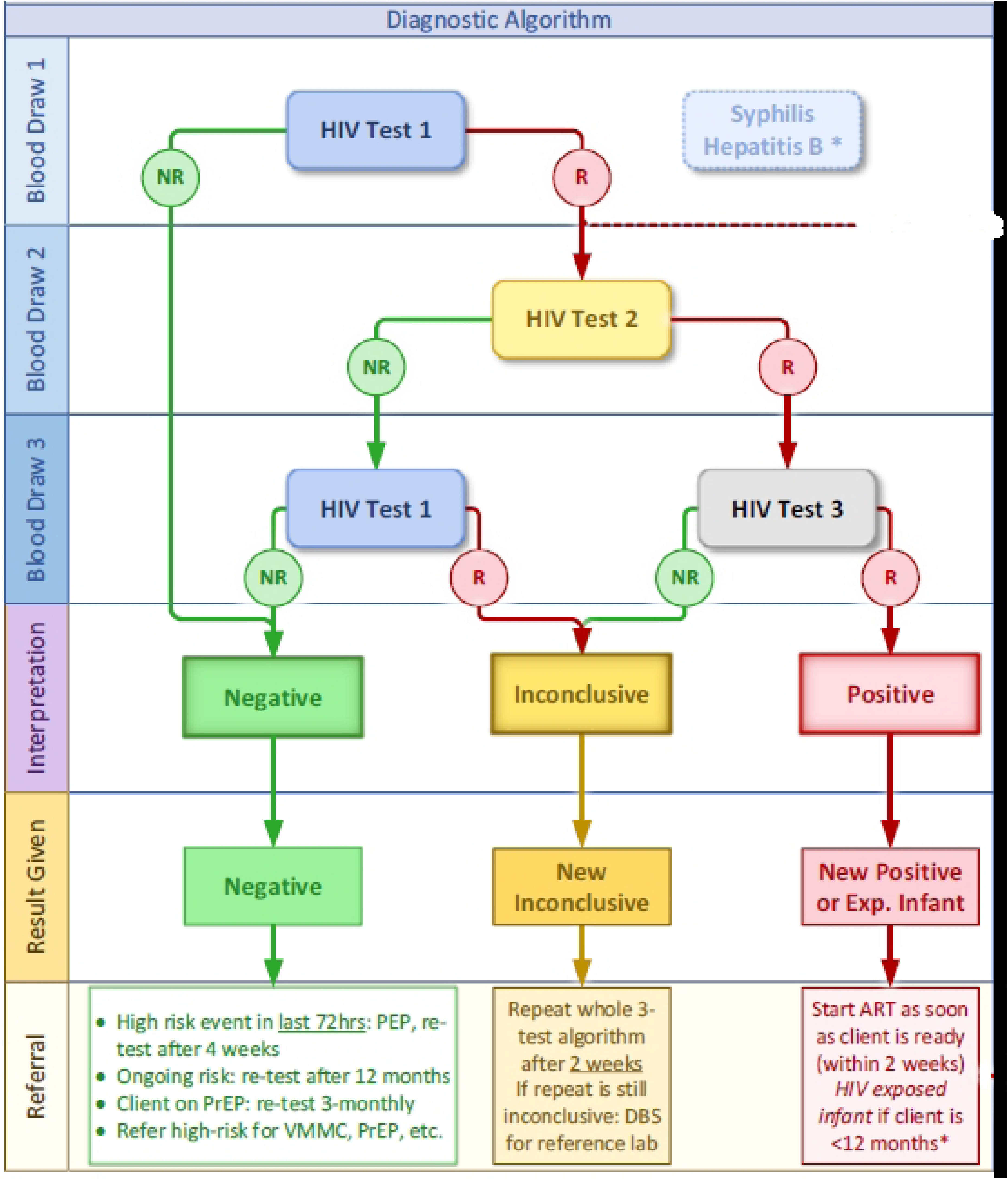
Three-test HIV algorithm adopted in Malawi since November 2022.

### Developing the Malawi ScanForm-based HTS M&E system

#### Overview of the ScanForm technology

The ScanForm M&E system consists of scannable paper registers, Android smartphones equipped with the ScanForm application, and automated reporting tools [16]. Health providers photograph the scannable paper registers using the ScanForm mobile app, and the images are uploaded automatically to a central server (**Fig 2**). Data are encrypted both at rest and in transit using Federal Information Processing Standards (FIPS)-compliant algorithms, and images are automatically deleted from the smartphones 90 days after upload. ScanForm uses computer vision algorithms and deep-learning models tuned to recognise locally prevalent handwritten styles. After the upload, data are extracted from the images into individual-level records with fields flagged for human verification if the optical character recognition (OCR) model is uncertain. As an additional safeguard, critical fields can be configured to always be shown for human verification, regardless of how certain the models are. In Malawi, this option is applied to visit dates and HIV test results. The verified individual-level data are subjected to validation checks. Detected validation errors are sent to facility Android smartphones equipped with the ScanForm application at the facility, where health providers correct the paper register and recapture the image for processing. Processed data are then automatically aggregated and presented as custom summary statistics and visualisations.

**Fig 2.**
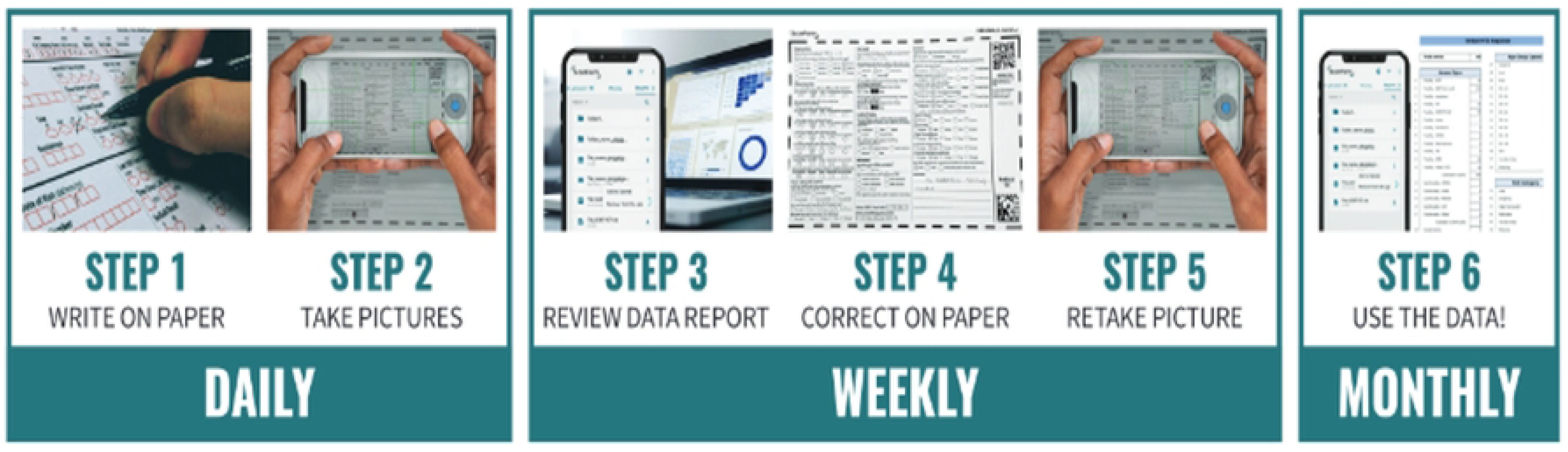
ScanForm process, from image capture to data utilisation.

Smartphones are secured with mobile device management bundled with ScanForm. This provides remote tracking and administration, disabling factory reset, monitoring of mobile data usage, and preventing use or installation of applications that are unrelated to management of health program data. The server can be locally hosted when an official national data centre is available.

ScanForm architecture supports integration with systems like DHIS2, OpenMRS, and OpenHIM. As of 2025, ScanForm has been used in various projects in Malawi, Kenya, Cameroon, Nigeria, Côte d’Ivoire, South Africa, Zambia, Benin, and Burkina Faso.

#### Designing Scannable Registers for the Malawi HTS Program

Previously, all HTS data were recorded in a single HTS register with personal identifying information (PII). Since most HTS clients in Malawi (97%) test negative for the first HIV test and require no follow-up, their PII, and all follow-up questions on subsequent HIV tests, recency, dried blood spot (DBS) samples, and antiretroviral therapy (ART), were unnecessary. Instead of developing one HTS register, which would result in almost 77% of paper in the register space being wasted, a multidisciplinary MoH, organisations offering HIV testing services, and the QED team developed two interlinked scannable registers: the initial HIV testing register and the HIV confirmatory register (**S1 file**). The initial register captures facility details, demographic information, HIV risk and testing history, T1 results, and tests for hepatitis B and syphilis as part of the triple elimination strategy. The confirmatory register, used only for T1-reactive clients, captures PII for follow-up by the facility (first and last name, phone number, residence), T2, T3, and repeat T1 results, final interpretation of the three HIV tests, HIV recency test results, DBS specimen IDs, and referrals for ART and other services. PII is recorded in a non-scannable area of the confirmatory register, to support follow-ups without digitisation. The initial and confirmatory registers contain 24 and 30 data elements per client, respectively.

#### Customising ScanForm technology for the Malawi HTS Program

ScanForm’s AI was benchmarked against the Malawian handwriting styles. Handwriting samples were collected from 1,471 Malawian healthcare workers using a standardised form containing all alphabetic letters, digits, and checkboxes. Each form was captured with a ScanForm-enabled smartphone and processed. Overall OCR accuracy, prior to human verification, was 98%, with 98.4% (112,447/114,261) for letters, 99.2% (43,460/43,829) for digits, and 100% (51,484/51,484) for checkboxes. This baseline performance was deemed acceptable by Malawi MoH to proceed with a pilot and scale-up.

#### Automated validation checks

Automated validation checks were developed for both registers to detect recording errors and HTS protocol deviations. Validation checks for recording errors included inaccurate, missing, or inconsistent values within the HTS encounter record, while validation checks for testing protocol violation referred to deviations from the three-test algorithm, HTS standard operating procedures and misinterpretation of results. A single HTS encounter could have multiple validation checks.

**Table 1** shows a subset of autogenerated validation check messages. Results of the validation checks were shared with facilities through smartphones and a web-based portal. HTS providers were expected to resolve the anomalies promptly, although corrections were allowed up to six months after the date of the HTS encounter (visit date). If a correction created another error, a new validation check message was generated.

**Table 1.** A subset of validation check messages generated by the ScanForm-based HTS M&E system.

| Validation check message |
| --- |
| [Recorded value] in 'Time Since Last HIV Test Quantity' should contain only numbers |
| 'ART Clinic Registration Number' should be filled out, because 'ART Referral Outcome' was 'Linked' |
| 'Age' cannot be zero |
| 'DBS Collected' should be 'Yes' because 'DBS ID' was filled out and 'RTRI Result' was 'Recent' |
| '[recorded date]' in 'Visit Date' should be less than or equal to '[recorded date]' in 'Recency Test - Kit Expiry Date' |
| 'Referral for ART' should be 'No' because 'Result Given to Client' was inconclusive |
| 'Referral for Re-Testing' should not be 'Confirmatory Re-Test' because there is no linked confirmatory record and HIV Test 1 is not positive |
| 'Visit Date' of '[recorded date]' is incorrect because it occurred before the project start date of '[recorded date]' |
| Access Point [Recorded access code] does not exist |
| All tests are either not done, missing, or invalid ('HIV Test 1 Result', 'Hepatitis', 'Syphilis') |
| Breastfeeding female cannot be an infant, so her last HIV test should not be 'Exposed Infant' |
| Could not find Link ID 'No' in the confirmatory register for this client with positive HIV Test 1 |
| DBS should be collected because 'Last HIV Test' was 'Inconclusive' and 'Result Given to Client' was 'Inconclusive Re-test' |
| Incorrect date in field 'Referral Visit Date': [recorded date] |
| Partner was present at session, but Partner Status was also specified as not present |
| Pregnant female cannot be an infant, so her last HIV test should not be 'Exposed Infant' |
| Recency test should not be done for clients under 13 years of age |
| Too many answers selected for 'Client Risk Category', only one should be chosen |
| Value is missing in 'HIV Test 1 - Kit Expiry Date' |

The QED technical support team used WhatsApp group chats with the HTS providers from each facility to share observations and provide remote support on data quality and HTS protocol violations. During scheduled supportive supervision visits, HTS supervisors from MoH and organisations supporting HTS services offered additional support to facilities.

#### Program monitoring and reporting

A web-based password-protected analytics portal with interactive dashboards, validation checks files and monthly program reports was developed and updated daily to support monitoring of program performance and data quality (**Fig 3**). The portal allowed data to be disaggregated by district, facility, sex, age group, reporting periods, and organisations providing HTS.

**Fig 3.**
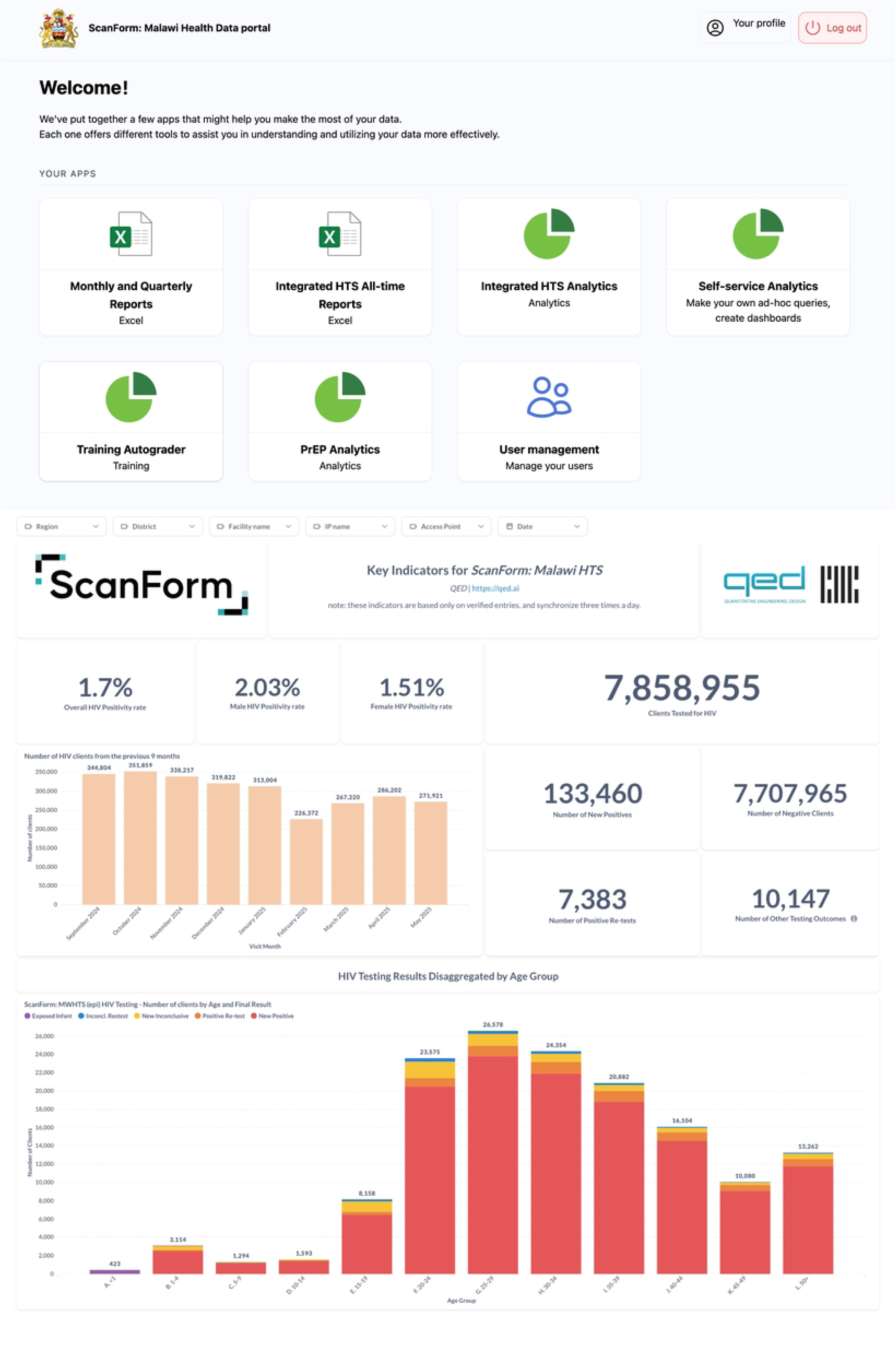
Malawi HIV Testing Services Data portal.

In addition to sharing data through the HTS portal, individual-level HTS data were transmitted daily to Malawi’s central data repository, while aggregated monthly reports transmitted to the DHAMIS and DHIS2 on the 5^th^ of every month, replacing manual data abstraction from paper registers. Individuals with HTS portal account received email alerts on the fifth of each month about the new aggregated monthly reports.

### Pilot and assessments

Nine facilities were selected purposively based on high client volumes and high HIV testing positivity to ensure sufficient data to evaluate the ScanForm M&E system and the three-test algorithm. The M&E system and the three-test algorithm were piloted between March and December 2021. To assess HTS providers’ experience with the new M&E system and the three-test algorithm, an independent organisation, not involved in the system development or implementation, conducted a survey and Focus Group Discussions (FGD) with HTS providers at the pilot sites between 27 May and 18 August, 2021 (**S2 file**).

### Site activation for national scale-up

Since the number of HTS facilities could increase during the scale-up period, making tracking coverage challenging, the Malawi MoH planned to implement the ScanForm-based M&E system and three-test algorithm in 867 ART facilities which accounted for 98% of the national HTS encounters. All HTS providers in these facilities received training on the use of the M&E tools before site activation. During activation, M&E officers and HTS program staff assessed facilities and, if necessary, re-oriented HTS providers. Once activated, facilities began using tools to record HTS encounters. Activation was conducted using a phased approach.

### Study outcomes and definitions

We assessed the following outcomes:

- M&E system coverage: Proportion of HTS encounters captured using the system by calendar months.
- Three-test algorithm coverage: Proportion of health facilities implementing the three-test algorithm by calendar months.
- Performance of the M&E system: (i) Number of validation check results: Number of validation checks results (recording errors and HTS protocol deviations) generated automatically from the first image submitted per register page, overall and by months since site activation. (ii) Error rate was calculated as the total number of recording errors and HTS protocol deviations divided by the number of data elements collected during the study period; (iii) data timeliness: Proportion of HTS encounters submitted no later than the fifth day of the following month after the client’s visit; (iv) Data completeness: Proportion of mandatory fields in HTS registers that were completed in the *final* HTS dataset.
- Implementation costs for the M&E system: Total financial resources required to implement the M&E system, including equipment, data plans, printing, and technical support.
- User experience: HTS providers’ feedback on usability and acceptability.

### Data analysis

Data for HTS encounters and validation check results were extracted from the server between 1 October 2025 and 30 November 2025 and analysed in R version 4.5.1 and Stata version 18. Descriptive statistics (counts and proportions) were used to summarise coverage of the M&E system and three-test algorithm, data timeliness, data completeness, implementation costs, and volume of validation check results. Validation check results were classified as “critical” if the issue affected monthly program summary statistics and required correction, or “non-critical” if the issue did not affect summary statistics and did not require data cleaning. To assess trends in validation check results, facilities with at least 24 months of follow-up after activation were included. Monthly counts of validation check results and error rates were calculated over the 24-month period since activation. Survey data and transcripts from focus group discussions (FGD) with HTS providers were analysed to assess user usability and acceptability.

### Ethical considerations

The Malawi National Health Sciences Research Committee reviewed and approved the protocol (Protocol number 23/12/4275). Healthcare workers involved in the qualitative assessment provided verbal informed consent, which was documented in research notes by the study assistants, and healthcare workers were assured that their decision to participate or not would not affect their employment. Informed consent from HTS clients was waived, as the study used programmatic data without personal identifiers.

## Results

### Coverage of the ScanForm M&E system and three-test HIV algorithm

By January 2023, 260 of 867 facilities (30%) had adopted both the M&E system and three-test algorithm. By September 2024, the implementation had scaled up to 853 (98%). The proportion of HTS encounters captured through the system rose from 56% in January 2023 to 98% by September 2024 (**Fig 4**). By April 2025, all 867 facilities implemented the ScanForm M&E system and the three-test algorithm. A total of 9,082,891 HTS encounters were collected through the system by September 2025; 761,248 from community testing services and 8,319,875 from facility-based services. Of the 8,319,875 HTS encounters, 153, 088 (1.8%) were positive.

**Fig 4.**
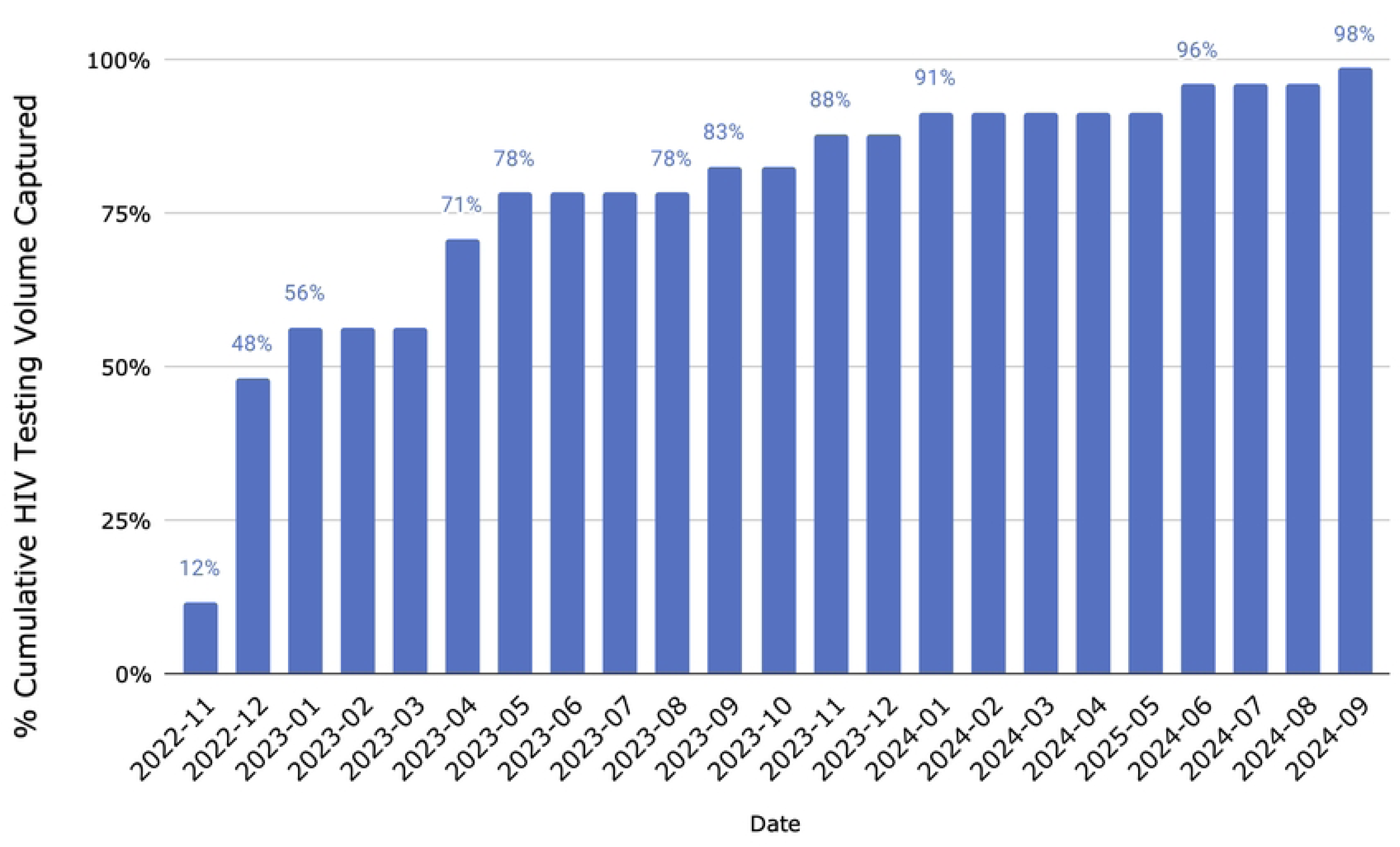
Volume of HIV testing encounters between November 2022 and September 2024 in Malawi.

### Performance of the M&E system

#### Automated validation check results for all facilities

A total of 1,412,133 validation check results were generated and sent to facilities, representing 0.6% error rate (1,412,133/245,238,057). Of these, 485,406 (34%) were classified as critical and 926,727 (66%) non-critical (**Table 2**). Of the 1,412,133 validation check results, 1,370,002 (97%) were recording errors and 42,131 (3%) HTS protocol deviations. Among the HTS protocol deviations, only two violation types were specific to the implementation of the three-test algorithm: 15,051 (36%) deviations from the HTS testing algorithm and 4,683 (11%) misclassifications of HIV test results.

**Table 2:**
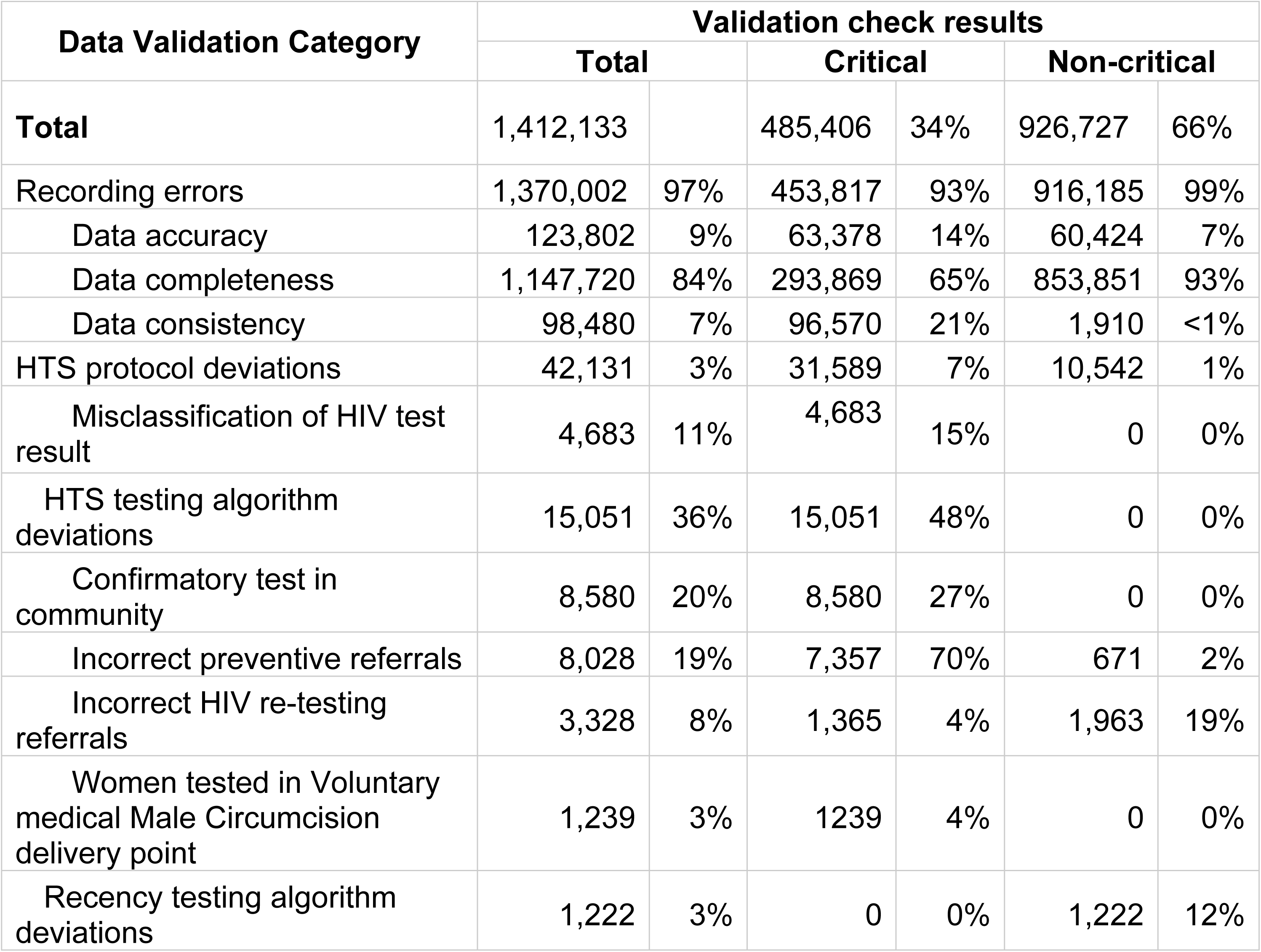
Validation check results generated by ScanForm HTS M&E system from November 2022 to September 2025 in Malawi.

| Data Validation Category | Validation check results |  |  |  |  |  |
| --- | --- | --- | --- | --- | --- | --- |
|  | Total |  | Critical |  | Non-critical |  |
| **Total** | 1,412,133 |  | 485,406 | 34% | 926,727 | 66% |
| Recording errors | 1,370,002 | 97% | 453,817 | 93% | 916,185 | 99% |
| Data accuracy | 123,802 | 9% | 63,378 | 14% | 60,424 | 7% |
| Data completeness | 1,147,720 | 84% | 293,869 | 65% | 853,851 | 93% |
| Data consistency | 98,480 | 7% | 96,570 | 21% | 1,910 | <1% |
| HTS protocol deviations | 42,131 | 3% | 31,589 | 7% | 10,542 | 1% |
| Misclassification of HIV test result | 4,683 | 11% | 4,683 | 15% | 0 | 0% |
| HTS testing algorithm deviations | 15,051 | 36% | 15,051 | 48% | 0 | 0% |
| Confirmatory test in community | 8,580 | 20% | 8,580 | 27% | 0 | 0% |
| Incorrect preventive referrals | 8,028 | 19% | 7,357 | 70% | 671 | 2% |
| Incorrect HIV re-testing referrals | 3,328 | 8% | 1,365 | 4% | 1,963 | 19% |
| Women tested in Voluntary medical Male Circumcision delivery point | 1,239 | 3% | 1239 | 4% | 0 | 0% |
| Recency testing algorithm deviations | 1,222 | 3% | 0 | 0% | 1,222 | 12% |

Among the 4,683 incorrectly specified results, 784 had potential clinical implications (**Fig 5**), while for the remaining 3,899 cases, the misclassification would not have altered the services provided to the client.

**Fig 5.**
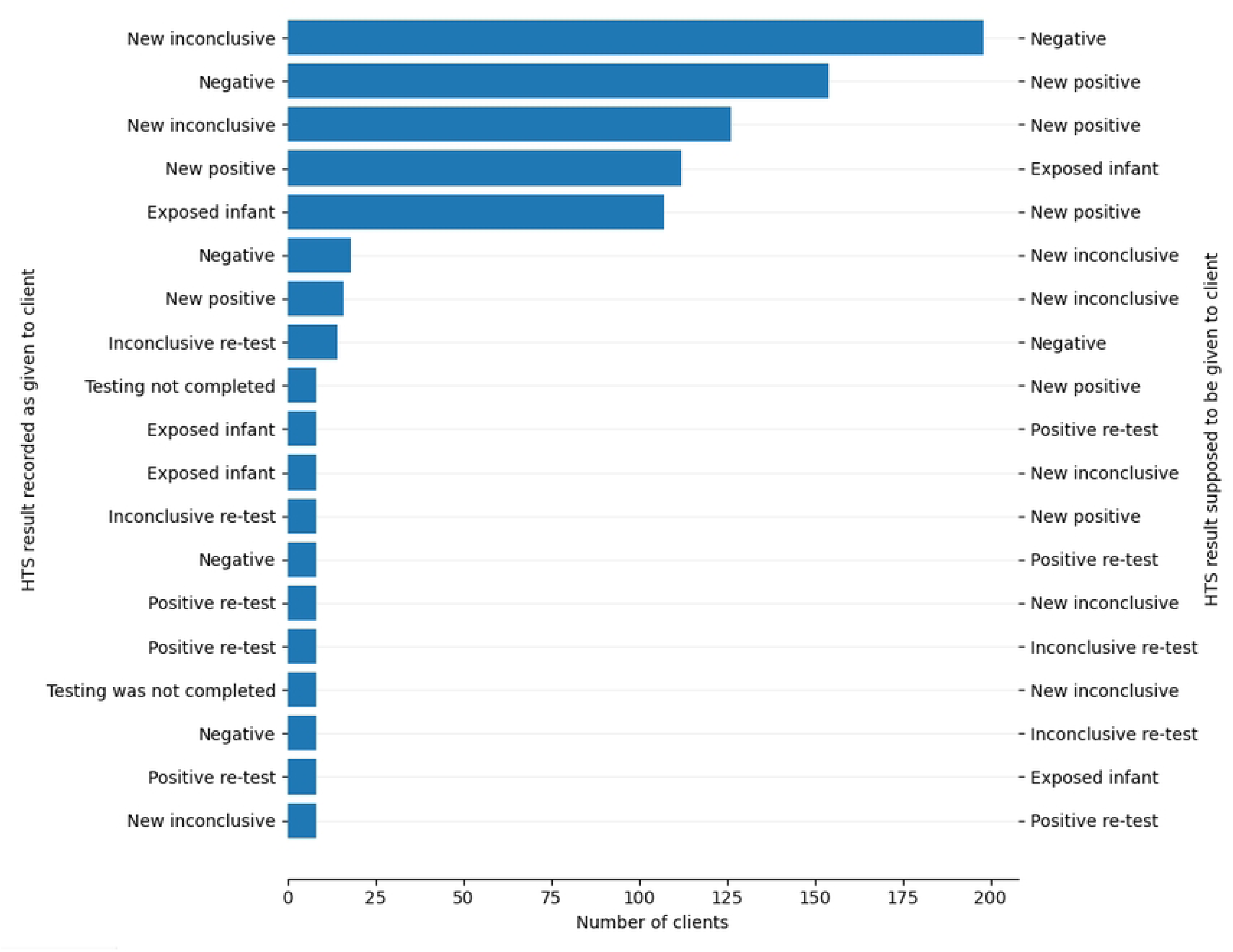
Misclassification of HIV test results generated by ScanForm HTS M&E system from November 2022 to September 2025 in Malawi.

As of 30 September 2025, 145,893 validation check results were not resolved. Of these, 130,240 (89%) were recording errors; 26,595 critical and 103,645 non-critical issues.

HTS protocol deviations were 15,653 (10%) which included 10,534 (67%) critical and 5119(33%) non-critical issues. Among testing protocol violations, 2 369 (15%) were testing algorithm deviations and 1,484 (9%) were misclassification of HIV test result.

#### Automated validation check results for facilities with at least 24 months of follow-up

A total of 469 facilities had at least 24 months of follow-up and were included in the longitudinal analysis of the validation check results. A total of 907,255 validation check results were analysed. The overall median number of validation check results per facility per month was 39 (mean 81). **Fig 6** shows HTS encounters, validation checks and error density by months since individual site activation. The number of HTS encounters increased gradually by 60% between the first and 24^th^ month of follow-up after site activation. During the first three months, validation checks declined by 32% and by 58% over the entire 24-month period. The error rate decreased from 1.6% to 0.4% between the first month and 12 months of follow-up, and remained stable at 0.4% thereafter.

**Fig 6.**
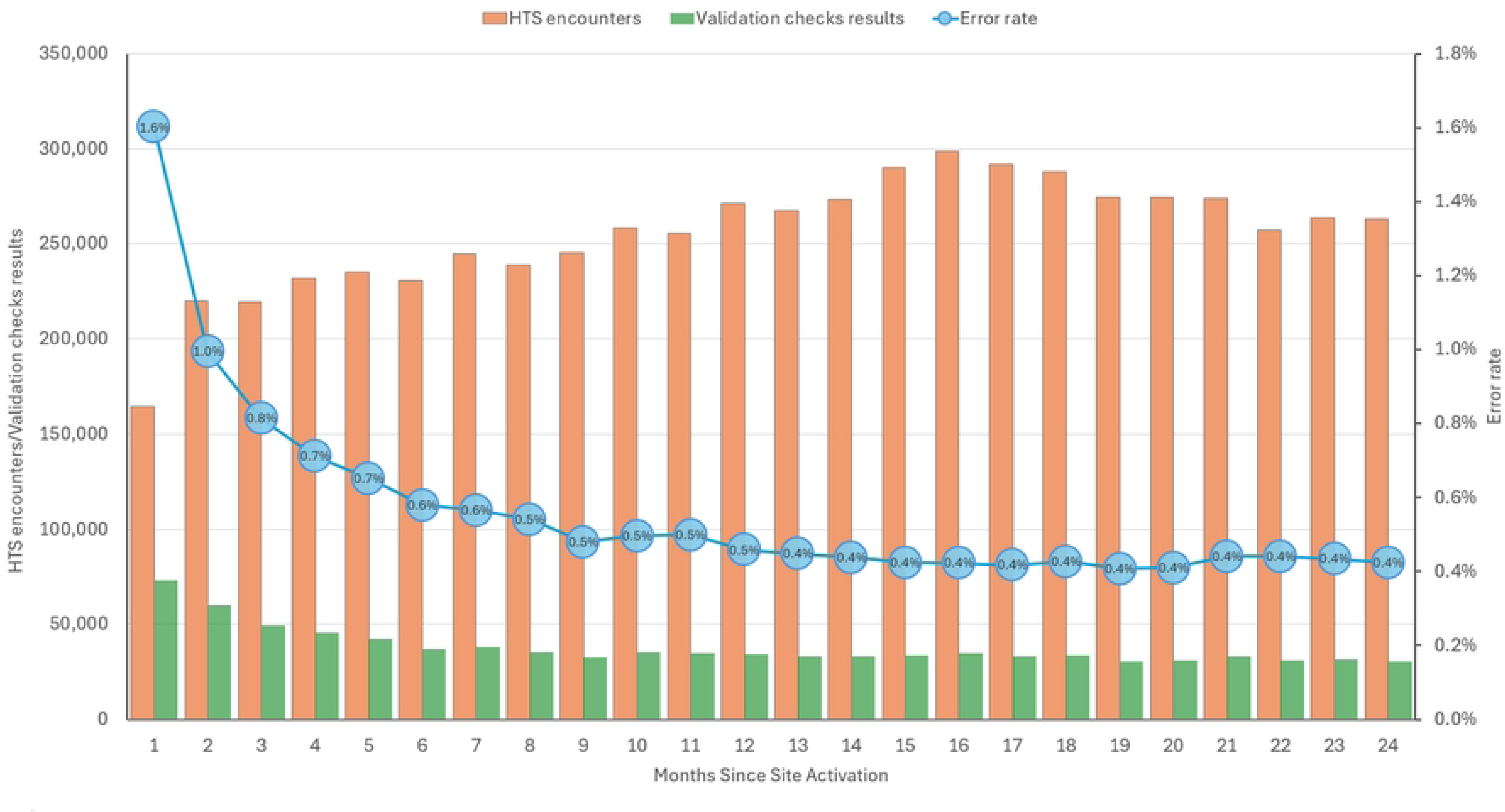
Trends in HTS encounters, validation check results, and error rate by Month Since site activation of ScanForm M&E system tool and three-test HIV algorithm in Malawi. *Note: The chart included only facilities with 24 full months of follow-up, using respective site activation month as the baseline. An HTS encounter could have multiple validation checks*.

#### HTS data timeliness

Approximately 94.6% of all HTS records were submitted on time (**Fig 7**). Submission rates were stable overall but declined to 86% between January and April 2025.

**Fig 7.**
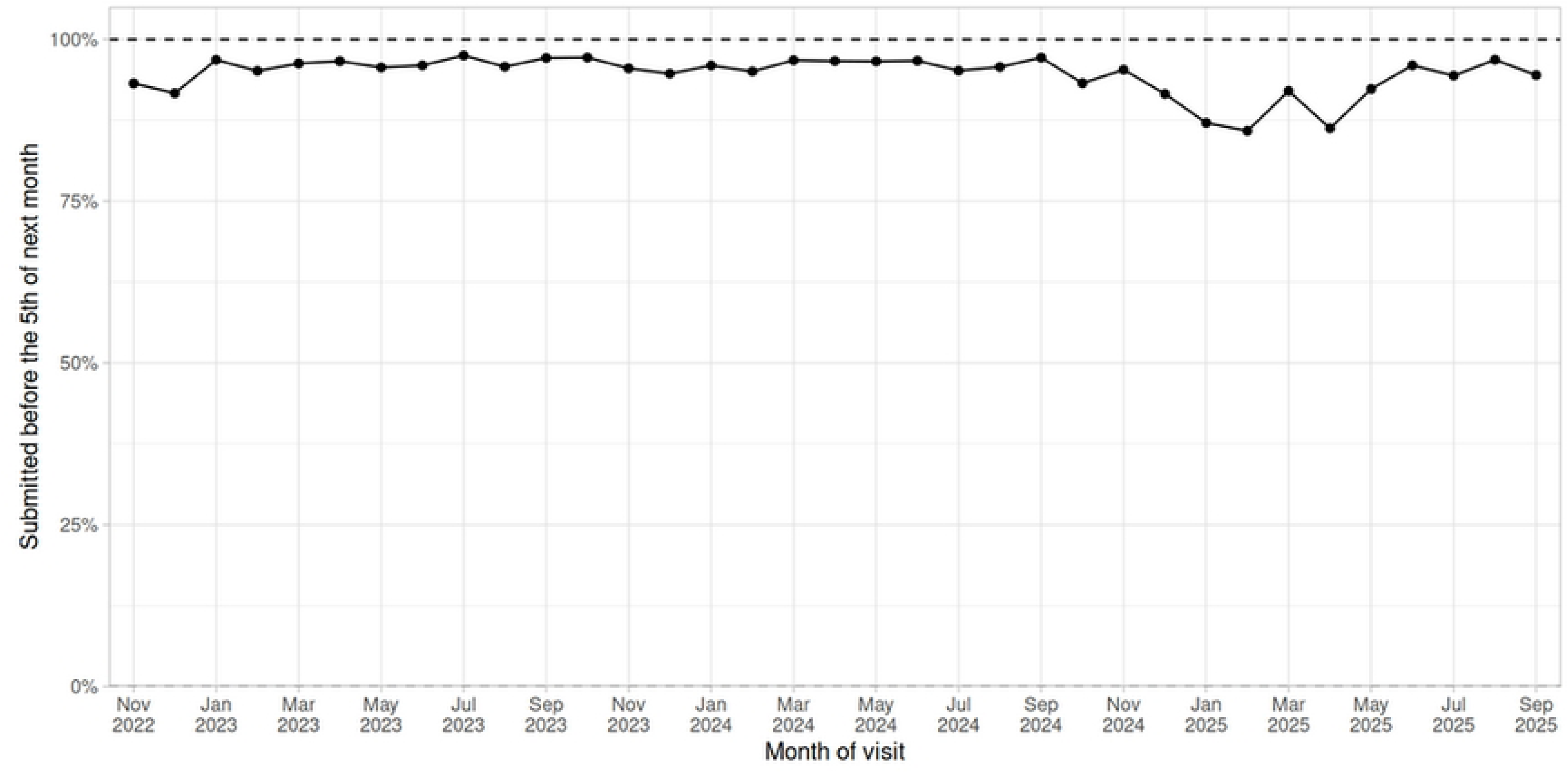
Proportion of HTS encounters submitted before the 5th of the following months after HTS client’s visit in Malawi.

#### HTS data completeness

Of the 9,082,891 HTS encounters, there were 141,308,486 mandatory data elements. Before data cleaning, 99.6% of the records were complete, which increased to 99.9% after data cleaning (**Table 3**). Data incompleteness was common among conditionally required fields (between 1% and 6%).

**Table 3.** Completion of mandatory fields in ScanForm M&E registers (Initial and Confirmatory) by HIV testing providers in Malawi, from November 2022 to September 2025.

|  | Mandatory Entries | Missing value (n, %) |  |
| --- | --- | --- | --- |
| Records | 9,082,891 |  |  |
| Total | 141,308,486 | 180,370 | 0.13% |
| Visit date <sup>β</sup> | 9,261,387 | 0 | 0.00% |
| Sex | 9,261,387 | 0 | 0.00% |
| Age | 9,261,387 | 4,278 | 0.05% |
| Last HIV test | 9,091,022 | 0 | 0.00% |
| Time since last HIV test <sup>‡</sup> | 7,363,913 | 92,482 | 1.26% |
| Ever taken ARV | 9,091,022 | 0 | 0.00% |
| Time since last ARV <sup>‡</sup> | 283,848 | 20,947 | 7.38% |
| HIV exposure category | 9,091,022 | 1,037 | 0.01% |
| HIV test 1 | 9,091,022 | 7 | 0.00% |
| Hepatis B | 9,091,022 | 423 | 0.00% |
| Syphilis | 9,091,022 | 566 | 0.01% |
| Partner Present | 9,091,022 | 191 | 0.00% |
| Partner HIV Status | 9,091,022 | 0 | 0.00% |
| Referral for testing | 9,261,387 | 0 | 0.00% |
| Retesting date <sup>‡</sup> | 3,844,088 | 34,414 | 0.90% |
| Link ID | 170,365 | 0 | 0.00% |
| Provider ID | 9,261,387 | 0 | 0.00% |
| Access point code | 9,261,387 | 0 | 0.00% |
| Test Two | 170,365 | 549 | 0.32% |
| Test three / Test 1 repeat | 170,365 | 590 | 0.35% |
| Final HIV result given | 170,365 | 87 | 0.05% |
| Results for rapid test for recent HIV infection | 170,365 | 0 | 0.00% |
| Dry blood sample collected | 170,365 | 0 | 0.00% |
| Referral for ART <sup>‡</sup> | 170,365 | 58 | 0.03% |
| Referral date <sup>‡</sup> | 157,219 | 24 | 0.02% |
| ART referral outcome <sup>‡</sup> | 170,365 | 9,333 | 5.48% |

<sup>‡</sup> Conditionally required fields, <sup>β</sup> Visit date field is from initial and confirmatory registers

**Table 4.** Implementation cost of ScanForm M&E system in Malawi.

|  | Total | Year 1 (Nov 2022-Oct 2023) | Year 2 (Nov 2023-Oct 2024) | Year 3 (Nov 2024-Sep 2025) |
| --- | --- | --- | --- | --- |
| Number of sites | 864 | 489 | 846 | 854 |
| Number of HTS encounters | 9,082,891 | 1,962,843 | 3,831,212 | 3,288,836 |
| **Cost components** |  |  |  |  |
| ScanForm Core Cost<sup>β</sup> | \$1,220,000.00 | \$660,000.00 | \$560,000.00 | \$300,500.00 |
| Printing of HTS registers | \$135,000.00 | \$67,500.00 | \$67,500.00 | \$67,500.00 |
| Smartphone | \$167,000.00 | \$135,000.00 | \$32,000.00 | \$32,000.00 |
| Data plans | \$60,000.00 | \$30,000.00 | \$30,000.00 | \$30,000.00 |
| Total | \$1,582,000.00 | \$892,500.00 | \$689,500.00 | \$430,000.00 |
| Average cost per site | \$1,831.02 | \$1,825.15 | \$815.01 | \$503.51 |
| Average cost per HTS record | \$0.17 | \$0.45 | \$0.18 | \$0.13 |

<sup>β</sup>Includes customization of ScanForm, designing HTS registers, development of reporting pipeline and technical support.

### Implementation costs of ScanForm M&E system

During the first three years of implementation, the total implementation cost was US$1.58 million, covering the core system (software customisation, designing scannable registers, data pipeline development, and technical support), printing of scannable HTS registers, smartphones, and data plans for mobile phones. Overall, the average implementation cost was $1,831 per site and $0.17 per HTS encounter. Implementation costs per site decreased from $1,825.15 in Year 1 to $815 in Year 2, and $503.51 in Year 3, while the cost per HTS record declined from $0.45 in Year 1 to $0.13 by Year 3.

### HTS Providers’ Experiences with ScanForm M&E system

Of the 42 HTS providers who participated in the survey, 95% (40/42) rated the AI-powered M&E system as a good or very good technology for managing HTS data. Most (95%) reported time savings, and all (100%) indicated faster data collection due to tick-box responses. In FGDs with 53 providers, participants highlighted benefits of the system, such as ease of use and reduced workload, as the system’s automated reporting replaced manual data compilation. One provider noted,

> *“It was very easy because every time when you work, you are supposed to write a report but in this case, once we fill the form and upload, we were done with the report.”*

Some participants noted challenges with small font sizes in the registers, which impeded documentation, and the absence of row numbers in the initial HTS register, which occasionally led to skipped lines. These issues were addressed in subsequent versions of the HTS register.

## Discussion

Malawi was one of the first countries to implement the 2019 WHO-recommended three-test algorithm for HIV diagnosis at a national scale. This study assessed the coverage and performance of ScanForm M&E system deployed to monitor HIV testing services quality assurance and to generate routine program reporting during the three-test algorithm scale-up. Scale-up of both interventions was rapid, achieving full national coverage within two years. The sustained reduction in error density suggests improvements in data recording quality and adherence to HIV testing protocols. Furthermore, high data completeness, consistent timely reporting, and the availability of automated reports demonstrated improved data quality and supported program oversight.

The ScanForm-based M&E system was designed to address the three well-documented barriers to EMRs adoption in SSA. First, infrastructure constraints, including inadequate equipment, poor internet connectivity, and power shortages [11–13] were addressed by allowing providers across multiple HTS delivery points share a single smartphone as individual devices were not required at each HTS delivery point. The smartphones had an offline-capable workflow that ensured continuous data capture without reliance on constant power or internet connectivity. Second, EMRS implementation is challenged by limited digital literacy and inadequate user support [17]. The ScanForm-based M&E system mitigated these barriers by using paper and pen familiar to healthcare workers, paired with a digitization process conducted using a simple smartphone camera. Additionally, QED provided sustained remote mentorship via phone. Finally, the system provided near-real-time individual level data within hours of images digitisation and reduced staff workload by automating routine report generation. Together, these features reduced technical and human resource barriers that typically limit large-scale implementation of digital systems.

The successful scale-up of the ScanForm M&E system alongside the three-test algorithm was mainly driven by Malawi’s robust HIV testing program and effective coordination mechanisms. In partnership with non-governmental organisations providing HTS, the Malawi government established clear policy direction and held regular meetings to align stakeholders, coordinate resources and monitor progress using the web-based analytics portal. Scaling a national HTS workforce to implement both the three-test algorithm and the M&E system required substantial financial resources, which were not available at once. A phased rollout was therefore implemented to allow gradual site activation while managing training and operational capacity within available resources. During the phased rollout, high-volume HTS sites were prioritised, using a cascade training model and providing on-site mentorship during site activation to maintain training quality. By combining strategic coordination, data-driven monitoring and a phased implementation approach, nationwide adoption was rapid while maintaining program fidelity.

The monthly recording errors and HTS protocol deviations decreased over time, despite increasing testing volumes. The decline could be likely attributed to immediate feedback on recording errors and HTS protocol deviations provided to HTS providers, combined with ongoing remote support. These mechanisms served as a continuous learning opportunity, where HTS providers became aware of recording errors and protocol violations, resolved them and prevented recurrent errors. Although quarterly supportive supervision visits may have contributed to the improved performance, their effect would be expected to occur later rather than immediately as observed in this study. Despite achieving a sustained reduction, an error rate of 0.4% persisted. This likely reflects two operational realities: (1) turnover of HTS providers at facilities, which require onboarding; and (2) the inherent potential for human error even among trained staff. To further reduce recording errors and deviation from the three-test algorithm, the ScanForm system is being upgraded to provide immediate real-time feedback at the point of image capture to allow users correct potential validation issues before the images are uploaded to the central server. Overall, these findings demonstrate that the digital model with routine digital validation can improve data quality, support provider learning, and offer a scalable model for quality assurance that remains effective as service scales up.

Timely and complete data are critical for effective program monitoring and decision - making. Over 95% of HTS records were submitted on time, and aggregated monthly reports were uploaded to DHIS2 before the MoH recommended 15^th^ of the following month. The drop to 86% between January and April was as a result of the US government stop-work order, which affected NGO-employed HTS providers. In addition to timely submissions, the ScanForm M&E system achieved an overall high individual level data completeness (99.8%). Data completeness for EMRs in SSA varies widely, with reported rates ranging from 27% to above 92% ([18–20] lower completeness is frequently associated with routine or under-resourced settings. While ScanForm system supports HIV testing services rather than HIV clinical care supported by EMRs, its data completeness result places it at the upper end of the spectrum reported for digital health data systems in the region. This suggests that its design is effective for ensuring timely and complete data capture within its intended scope.

The ScanForm-based M&E system was associated with reductions in per-site costs over three years. When fully deployed across all sites in Malawi, the average cost per site decreased from US$1,825.15 in year 1 to US$503.51 in year 3. When considered against Malawi’s HIV testing volume annually, the Year 2 and Year 3 costs translate to about US$0.18 and US$0.13 per record, respectively, a small fraction of the average facility-based HIV testing per encounter (US$ 2.85, including M&E and data audit costs) in MalawiClick or tap here to enter text. [22]. As foreign funding declines and national budgets tighten [23], some traditional methods of data collection are unsustainable. AI-based solutions offer innovative approaches to sustain a high-quality, accountable, and data-driven HIV response.

## Limitations of the study

First, the performance during the pilot may be overestimated because HTS providers at high-volume, predominately urban facilities may not be representative of the broader range of HTS providers in Malawi, as these sites may have greater capacity than smaller, more remote facilities. Second, recording errors may have been misclassified as testing protocol deviations, especially when HTS providers followed the correct procedures, but recorded different HIV test results in the register. Fourth, observed declines in recording errors and HTS protocol violations could be influenced by staff turnover and changes in testing volumes, rather than solely reflecting improvements in provider skills. Despite these limitations, the study offers valuable practical insights into implementation of an AI-powered M&E system when rolling out new HIV testing guidelines in resource-limited settings.

## Conclusion

The AI-powered M&E system successfully supported monitoring the scale-up of the three-test algorithm and generated timely routine program reports. The system overcame infrastructure and technical limitations of traditional EMRs, while providing reliable individual-level data to strengthen program oversight. Sustained reduction in error rate indicates that the system improved data quality and adherence to the three-test algorithm protocols. Implementation of the system was also associated with cost reductions over time. AI-powered M&E systems can provide reliable, timely and cost-effective data to support large-scale HIV program implementation in resource-constrained settings.

## Data Availability

Data cannot be shared publicly because of the Malawi data policy. Data are available from the Directorate of HIV, STI and Viral Hepatitis, Ministry of Health, Lilongwe, Malawi (contact via) for researchers who meet the criteria for access to confidential data.

## Acknowledgements

The authors thank the staff who collected data using the ScanForm-based HTS registers and the following individuals for their contributions to the development and implementation of the three-test algorithm and the ScanForm-based M&E system: Dr. Rose Nyirenda, Anna Drabko, Jakub Pieszczek, Bartosz Wojno, Bartosz Smoczynski, Piotr Bakalarski, Adam Furtak, Michał Banaszkiewicz, Wiktor Garbarek, Anna Kramarska, Oskar Tołkacz, Jan Stefaniak, Piotr Wiszniewski, and Gracedel Palma. We also acknowledge the Global Fund for supporting the ScanForm-based M&E system and the U.S. President’s Emergency Plan for AIDS Relief (PEPFAR) for supporting HIV testing services.

TC and JWI-E were supported by the Gates Foundation (INV-005576). Under the grant conditions of the Gates Foundation, a Creative Commons Attribution 4.0 License has already been assigned to the Author Accepted Manuscript version that might arise from this submission.

The findings and conclusions of this study do not represent the official views of the funding agencies.

## Supporting information

*S1 file. Malawi HTS initial and confirmatory registers*

*S2 file. Survey questionnaire and Focus Group Discussion guide*

